# Effect of Low-intensity Transcranial Focused Ultrasound Stimulation on Neuropathic Pain: Protocol for a Randomised, Single-blind, Placebo-controlled, Three-arm, Parallel-group Pilot Trial (TUS-P-02 Study)

**DOI:** 10.1101/2025.11.05.25338279

**Authors:** Nobuhiko Mori, Yuhei Hoshikuma, Koichi Hosomi, Akihiro Yamamoto, Takeshi Shimizu, Hui Ming Khoo, Naoki Tani, Satoru Oshino, Haruhiko Kishima

**Affiliations:** Department of Neurosurgery, The University of Osaka Graduate School of Medicine, Suita, Osaka, Japan

**Keywords:** transcranial focused ultrasound stimulation, non-invasive brain stimulation, neuropathic pain, protocol, randomised controlled trial

## Abstract

**Introduction:** Neuropathic pain (NP) remains difficult to manage because conventional pharmacological therapies often have limited efficacy and intolerable side effects. Non-invasive neuromodulation techniques have emerged as potential alternatives with fewer adverse effects. Meta-analyses of randomised controlled trials suggest that high-frequency repetitive transcranial magnetic stimulation over the primary motor cortex has analgesic effects; however, its efficacy is modest and restricted by the inability to stimulate deeper brain regions. Low-intensity transcranial focused ultrasound stimulation (TUS) is a novel approach that enables precise targeting of deep brain structures and modulation of neural activity. Although promising in preclinical and preliminary human studies, its long-term therapeutic efficacy for NP remains unknown. This trial aims to evaluate the therapeutic effects and safety of repeated TUS sessions in patients with NP.

**Methods and analysis:** This randomised, participant-blinded, placebo-controlled, three-arm, parallel-group clinical trial will enrol 39 participants with NP (pain duration ≥ 3 months; pain intensity ≥ 4 on the Numerical Rating Scale). Participants will be randomly allocated to one of three groups: (1) TUS targeting the primary motor cortex, (2) TUS targeting the posterior superior insula, or (3) sham stimulation. Each participant will undergo weekly sessions for eight weeks. The primary outcome is change in weekly average pain intensity scores recorded in a pain diary. Secondary outcomes include additional pain scales, quality of life measures, psychological assessments, quantitative sensory testing, motor cortical excitability, and adverse events.

**Ethics and dissemination:** The protocol has been approved by the University of Osaka Clinical Research Review Board. Written informed consent will be obtained from all participants. Findings will be disseminated via scientific presentations and peer-reviewed publications.

**Trial registration number:** jRCTs052240227

**STRENGTHS AND LIMITATIONS OF THIS STUDY:**

⇒ This is the first randomised, participant-blinded, placebo-controlled clinical trial evaluating the long-term effects of transcranial focused ultrasound stimulation (TUS) for neuropathic pain.
⇒ TUS is a novel, non-invasive technique that can precisely target deep brain areas using a navigation system, offering the potential to modulate brain function.
⇒ The single-blind design (participants only) may introduce a risk of performance bias, as operators cannot be blinded owing to technical constraints.
⇒ The small sample size may limit statistical power to detect subtle between-group differences.

## INTRODUCTION

Neuropathic pain (NP) is defined by the International Association for the Study of Pain as “pain caused by a lesion or disease of the somatosensory nervous system” [1]. It often persists after the original injury has healed and is typically resistant to standard pharmacological treatments. Common forms include postherpetic neuralgia, painful diabetic neuropathy, phantom limb pain, spinal cord injury-related pain, and central post-stroke pain. A nationwide Japanese epidemiological survey estimated the prevalence of NP at 3.2%, corresponding to approximately 3.5 million adults in Japan [2]. Despite available therapies, many patients report inadequate pain relief, and adverse effects frequently limit long-term adherence. First-line agents such as gabapentinoids, tricyclic antidepressants, and serotonin-norepinephrine reuptake inhibitors are recommended, but their effectiveness remains limited, with numbers needed to treat ranging from 4.6 to 8.9 [3]. Repetitive transcranial magnetic stimulation (rTMS), a non-invasive neuromodulation technique that targets the primary motor cortex (M1), has been investigated as an alternative. Systematic reviews and meta-analyses suggest that rTMS provides analgesic benefits [3–5], although the efficacy remains modest, with a number needed to treat of 4.2. In our previous clinical trials, rTMS showed limited benefit for pain overall, and subgroup analyses revealed particularly reduced effects for lower limb pain, likely because rTMS cannot adequately stimulate deeper cortical regions such as the M1 foot area [6–8]. Because the M1 foot region and many other pain-related structures are located deep within the brain, further development of non-invasive therapeutic strategies capable of modulating these targets is needed.

In recent years, focused ultrasound has been increasingly developed and applied for a variety of purposes. This technology delivers ultrasound at frequencies above 20 kHz through the skull and can be directed to a localised brain region, allowing intervention in deep brain structures. Magnetic resonance-guided focused ultrasound for thalamic thermal ablation is already used in clinical practice for the treatment of tremors, while ongoing studies are exploring its role in microbubble-mediated opening of the blood–brain barrier. More recently, low-intensity transcranial focused ultrasound stimulation (TUS), which operates at intensities below 100 W/cm^2^, has emerged as a non-invasive technique to modulate neural activity from outside the skull. Over the past two decades, animal studies have shown that TUS can alter motor evoked potentials (MEPs) and regional cerebral blood flow [9]. Since its first application in humans in 2013, TUS has been reported to shorten reaction times in motor tasks, modulate somatosensory and visual evoked potentials, and influence perception. The proposed mechanism involves ultrasonic acoustic pressure acting on ion channels through mechanoreceptors, thereby modulating neuronal activity [10].

TUS is a novel technique capable of exerting localised effects on deep brain structures without surgery. In contrast, rTMS can stimulate only superficial cortical regions when applied focally, and attempts to target deeper areas inevitably result in diffuse activation. To date, no rTMS device has been developed that allows precise stimulation of deep brain structures. Potential therapeutic targets for NP include the M1 foot area, insula, cingulate gyrus, and thalamic nuclei. Although rTMS has been unable to adequately stimulate these regions, TUS may provide this capability. Thus far, only two studies of TUS in the context of pain have been conducted in healthy participants [11, 12], and one single-arm study has been reported in patients with NP [13]. We previously performed a sham-controlled, randomised, crossover feasibility study of TUS in patients with NP (jRCTs052230116). However, to the best of our knowledge, no randomised controlled trial has yet evaluated the clinical efficacy of TUS for NP. For clinical translation, the so-called offline effects—sustained aftereffects that persist beyond the stimulation period—are considered essential. As with rTMS, repeated sessions are likely required to achieve meaningful therapeutic benefits, rather than a single session.

Given the exploratory nature of this preliminary study, we selected two stimulation sites—M1 and the posterior superior insular cortex (PSI)—to assess their potential therapeutic effects. M1 was chosen based on extensive evidence from prior clinical studies using rTMS [3–5], whereas the PSI was selected because it is a major projection site of the spinothalamic tract and plays a key role in pain perception [14]. Furthermore, previous reports have shown that deep brain stimulation of the PSI can alleviate pain [15]. The aim of this study is to investigate the efficacy and safety of weekly TUS administered over eight weeks in patients with NP.

## METHODS AND ANALYSIS

### Trial design

This study is a randomised, single-blind (participant-blinded), placebo-controlled, parallel-group clinical trial conducted at the University of Osaka Hospital in Japan. Because pain is highly susceptible to placebo effects, a sham stimulation group was included as the control group to evaluate the superiority of active stimulation. A parallel-group design was selected instead of a crossover design owing to concerns about potential carryover effects and the risk of unblinding. To further explore potential differences in efficacy, two active stimulation sites were designated.

This study is an investigator-initiated clinical trial. Trial monitoring and auditing will be conducted by an independent academic research organisation (Department of Medical Innovation, the University of Osaka Hospital). Monitors will perform oversight in accordance with the monitoring plan to ensure data reliability and participant protection, verifying compliance with applicable laws, regulations, and the trial protocol, and will prepare monitoring reports. Data will be captured using an electronic data capture system (REDCap; Vanderbilt University, USA), which minimises input error, duplication, and missing data while ensuring security and transparency. The study was registered with the Japan Registry of Clinical Trials (https://jrct.mhlw.go.jp/en-top; jRCTs052240227) on 27 December 2024. The first patient was enrolled on 30 April 2025, and twelve patients had been recruited as of 5 November 2025. This protocol is based on Version 1.3, dated 25 March 2025.

### Participants

We will enrol patients aged 18 years or older with NP who meet the following inclusion criteria: (1) NP persisting for more than three months; (2) pain intensity score ≥ 4 on the Numerical Rating Scale (NRS) at screening; and (3) written informed consent to participate in this study. The definition of NP follows the International Association for the Study of Pain terminology [1], adopting probable or definite NP according to the Neuropathic Pain Grading System [16]. Exclusion criteria are as follows: (1) dementia with a Mini-Mental State Examination (MMSE) score ≤ 23; (2) severe aphasia or cognitive dysfunction; (3) serious psychiatric disorder; (4) history of epileptic seizures; (5) use of an implantable stimulator such as a cardiac pacemaker, except for implantable spinal cord stimulators; (6) metallic implants in the head, except for titanium products; (7) use of an implantable drug delivery system or implantable ventricular assist device; (8) pregnancy; (9) inability to complete the assessment questionnaires; (10) participation in any other clinical trial within three months prior to providing consent; and (11) any other condition deemed inappropriate for inclusion by the investigators. Participants will be recruited from outpatients of the Department of Neurosurgery at the University of Osaka Hospital.

Participants will be withdrawn from the study if any of the following criteria are met after enrolment: (1) the investigator determines that continuing the study poses an unacceptable risk owing to the occurrence of an adverse event; (2) the participant withdraws consent or requests to discontinue participation; (3) it is discovered that the participant does not meet the eligibility criteria; (4) the participant is unable to complete the required assessments or visits as a result of personal circumstances, such as relocation; or (5) the investigator determines, for any other reason, that discontinuation of the intervention is appropriate.

### Randomisation and blinding

Participants will be randomly allocated in a 1:1:1 ratio to one of three groups: (1) active stimulation targeting M1, (2) active stimulation targeting PSI, or (3) sham stimulation. Because the primary outcomes rely on participant-reported subjective assessments, this study adopts a single-blind design in which participants remain blinded to group allocation throughout the trial. Blinding of investigators delivering the intervention is not feasible with the current device. As this is a preliminary study in which most outcomes are patient-reported, investigators are not blinded.

Randomisation will be conducted using a stratified permuted block method, with the underlying cause of NP (central or peripheral origin) as the stratification factor. The allocation table and procedure will be generated by an independent technician who is not involved in patient enrolment or outcome assessment. Assignment will be managed through the REDCap randomisation module, which enhances secure handling of allocation information using password protection.

### Trial schedule

All patients who provide consent are assessed for eligibility by neurosurgeons specialising in pain management. Eligible participants will be randomly allocated to one of three groups and undergo baseline evaluations, including demographic and clinical characteristics. The intervention is administered once a week for eight weeks (Figure 1). This schedule was determined based on previous rTMS studies, which showed that intermittent stimulation—such as weekly sessions—provides significant pain relief between 4–8 weeks or sessions, after which the effect tends to plateau [17–20]. Participants will be asked to record a pain diary from one week prior to the intervention until one week after completion at week 8. The NRS for pain and paraesthesia (*shibire* in Japanese), along with the Short-Form McGill Pain Questionnaire 2 (SF-MPQ-2) [21], will be assessed at baseline, immediately after each intervention, and at follow-up visits. Additional measures—including the Brief Pain Inventory (BPI) [22], EuroQol 5 Dimensions 5 Level (EQ-5D-5L) [23], Hospital Anxiety and Depression Scale (HADS) [24], Pain Catastrophizing Scale (PCS) [25, 26], Pain Self-Efficacy Questionnaire (PSEQ) [27], MMSE, quantitative sensory testing (QST), motor cortical excitability, and the *Shibire* Questionnaire (validated by our group; manuscript in preparation) [28]—will be assessed at baseline and immediately after the final intervention at week 8. At the end of the study, Patient Global Impression of Change (PGIC) and blinding assessments will be collected. Adverse events will be monitored continuously from the start of the intervention until follow-up visits at weeks 12 and 16 or until study discontinuation. The detailed schedule is presented in Table 1.

**Figure 1.**
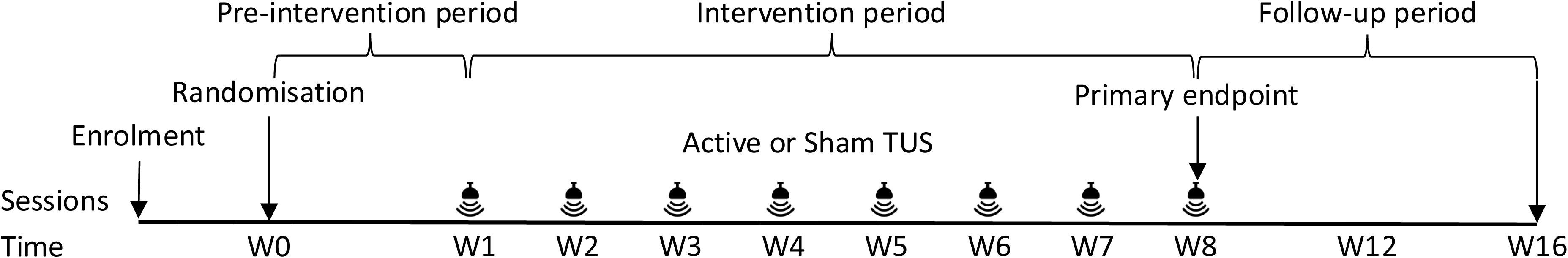
Trial schedule. TUS, transcranial focused ultrasound stimulation; W, week.

**Table 1.**
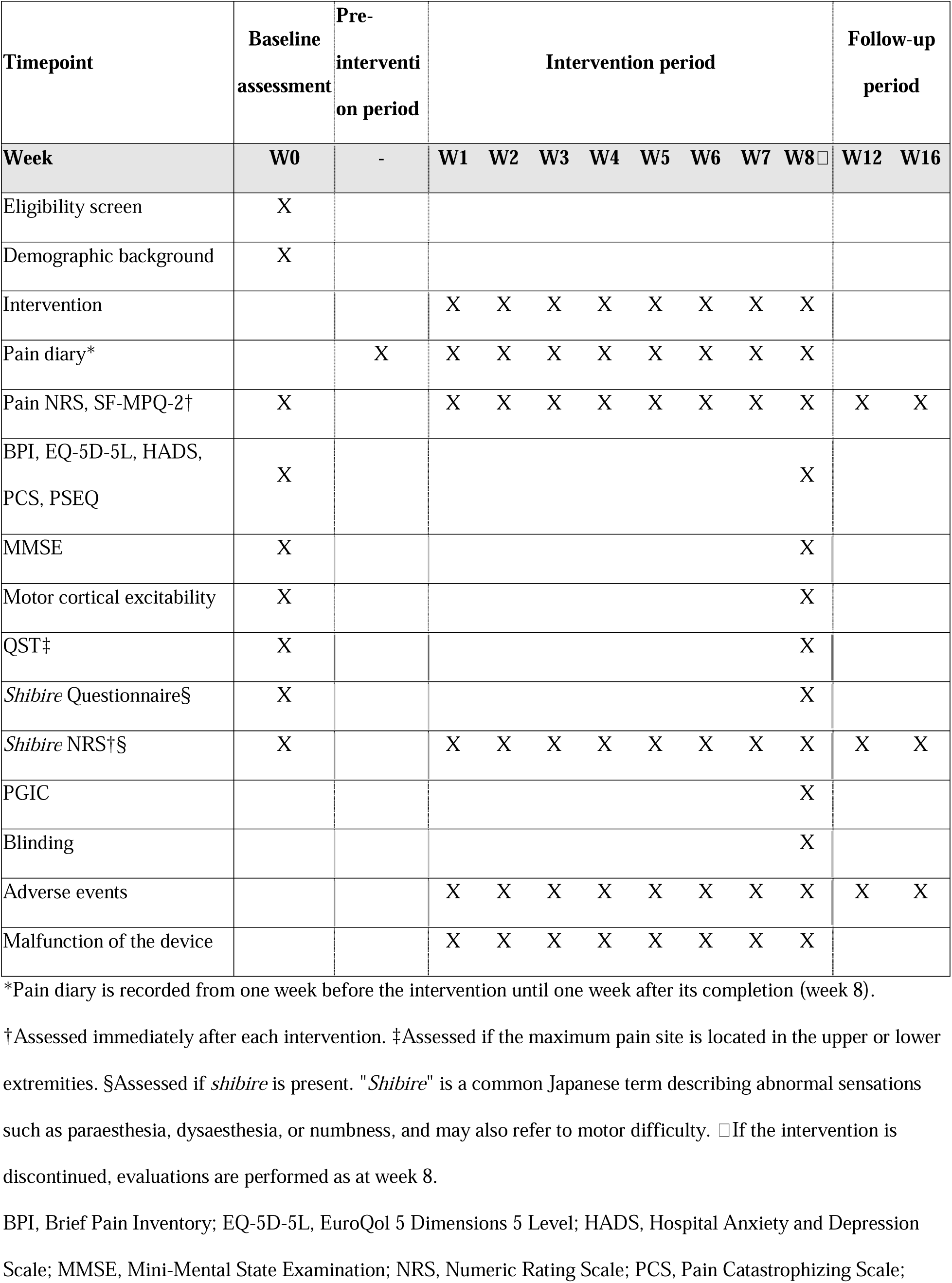
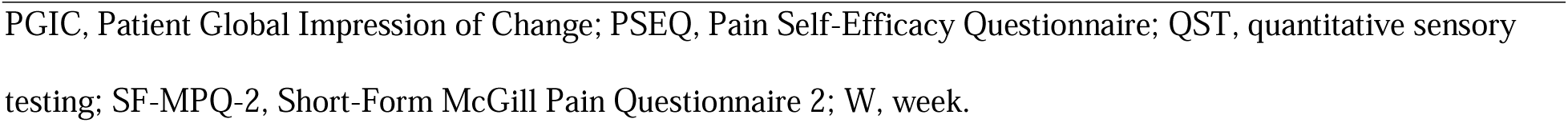
Schedule of enrolment, intervention, and assessment.

| Timepoint | Baseline assessment | Pre-intervention period | Intervention period |  |  |  |  |  |  |  | Follow-up period |  |
| --- | --- | --- | --- | --- | --- | --- | --- | --- | --- | --- | --- | --- |
| Week | W0 | - | W1 | W2 | W3 | W4 | W5 | W6 | W7 | W8□ | W12 | W16 |
| Eligibility screen | X |  |  |  |  |  |  |  |  |  |  |  |
| Demographic background | X |  |  |  |  |  |  |  |  |  |  |  |
| Intervention |  |  | X | X | X | X | X | X | X | X |  |  |
| Pain diary* |  | X | X | X | X | X | X | X | X | X |  |  |
| Pain NRS, SF-MPQ-2† | X |  | X | X | X | X | X | X | X | X | X | X |
| BPI, EQ-5D-5L, HADS, PCS, PSEQ | X |  |  |  |  |  |  |  |  | X |  |  |
| MMSE | X |  |  |  |  |  |  |  |  | X |  |  |
| Motor cortical excitability | X |  |  |  |  |  |  |  |  | X |  |  |
| QST‡ | X |  |  |  |  |  |  |  |  | X |  |  |
| Shibire Questionnaire§ | X |  |  |  |  |  |  |  |  | X |  |  |
| Shibire NRS†§ | X |  | X | X | X | X | X | X | X | X | X | X |
| PGIC |  |  |  |  |  |  |  |  |  | X |  |  |
| Blinding |  |  |  |  |  |  |  |  |  | X |  |  |
| Adverse events |  |  | X | X | X | X | X | X | X | X | X | X |
| Malfunction of the device |  |  | X | X | X | X | X | X | X | X |  |  |

\*Pain diary is recorded from one week before the intervention until one week after its completion (week 8).
†Assessed immediately after each intervention. ‡Assessed if the maximum pain site is located in the upper or lower extremities. §Assessed if *shibire* is present. "*Shibire*" is a common Japanese term describing abnormal sensations such as paraesthesia, dysaesthesia, or numbness, and may also refer to motor difficulty. □If the intervention is discontinued, evaluations are performed as at week 8.
BPI, Brief Pain Inventory; EQ-5D-5L, EuroQol 5 Dimensions 5 Level; HADS, Hospital Anxiety and Depression Scale; MMSE, Mini-Mental State Examination; NRS, Numeric Rating Scale; PCS, Pain Catastrophizing Scale;
PGIC, Patient Global Impression of Change; PSEQ, Pain Self-Efficacy Questionnaire; QST, quantitative sensory testing; SF-MPQ-2, Short-Form McGill Pain Questionnaire 2; W, week.

### Sample size estimation

As no previous clinical trials have examined the effects of repeated sessions of TUS in patients with chronic pain, the sample size calculation was based on a prior clinical trial of rTMS for NP [8]. In that study, the mean reduction in pain score immediately after the final intervention was 34.8 ± 15.2 in the active stimulation group (n = 15). Assuming that the sham group would experience half of this effect (mean reduction of 17.4), and that both active TUS groups in our study would yield the same mean reduction (34.8), the standard deviation was set at 15.2 for all groups. Based on these assumptions, a one-way analysis of variance with an alpha of 0.05 and 80% power to detect differences among the three groups indicated that 37 participants are required. Accounting for a 5% dropout rate, the final target sample size was set at 39 (13 per group).

### Interventions

TUS will be administered using the NeuroFUS system (Sonic Concepts, Bothell, WA, USA), which consists of a four-element annular array transducer (CTX-500, Sonic Concepts) with a diameter of 60 mm and a fundamental frequency of 500 kHz. The transducer is powered by a programmable radiofrequency amplifier (TPO-203, Sonic Concepts), which controls the phasing of the four elements to adjust the sonication depth. A neuro-navigation system (Brainsight; Rogue Research Inc., Montreal, QC, Canada) will be employed to identify the stimulation site and to precisely monitor and maintain the position and orientation of the transducer throughout all treatment sessions.

The stimulation target will be either M1, corresponding to the most painful region, or the PSI, both located contralateral to the pain. When the M1 hand area is selected, the motor hotspot identified through cortical excitability measurements using TMS will be used. The PSI will be localised on individual three-dimensional magnetic resonance imaging in the navigation system according to the previously reported “quadrant-within-a-quadrant” method [29]. For all other targets, stimulation sites will be determined on the individual magnetic resonance image with reference to the Montreal Neurological Institute space standard atlas integrated into the navigation system. Because the TUS device achieves optimal efficiency at a focal depth of 48.5 mm, an ultrasound gel pad of appropriate thickness (Echo gel pad, Yasojima, Kobe, Japan) will be placed between the transducer and the scalp to adjust the distance from the transducer surface to the stimulation target to within 43–54 mm. Stimulation parameters will be as follows: pulse duration, 10 ms; pulse repetition interval, 100 ms (10 Hz); pulse train duration, 10 s; pulse train repetition interval, 30 s; duty cycle, 10%; number of trains, 20; total stimulation duration, 580 s; spatial peak pulse average intensity in free water, 30 W/cm^2^. The mechanical index, estimated using the TUS Calculator (https://www.socsci.ru.nl/fusinitiative/tuscalculator/) at a focus depth of 43 mm, was 1.32. The maximum temperature rise under these stimulation parameters was calculated to be 1.33 °C, based on the estimated maximum rise reported by Osaka et al. using the same TUS device, intensity, and pulse repetition interval [30]. As the increase remained below 2 °C and the intervention time was less than 10 min, the thermal dose was estimated to be below 0.25 CEM43. The thermal index for the skull was calculated as 1.9. These values are within the safety thresholds recommended by the recent International Consortium for Transcranial Ultrasonic Stimulation Safety and Standards consensus on TUS safety [31].

Treatment will be administered with the participant seated comfortably in a reclining chair. Ultrasound gel will be applied between the scalp, gel pad, and transducer, with all visible air bubbles carefully removed. The transducer will then be affixed to the scalp via a gel pad to ensure complete contact. To maintain blinding, participants will not be shown the TUS device interface, and white noise will be delivered through earphones during stimulation to mask any device sounds. All interventions will be performed under the supervision of investigators who are also neurosurgeons specialised in neuromodulation. In the sham stimulation group, the transducer will be placed over M1, but no ultrasound will be emitted (0 W output), with the same device setup and procedures as in the active stimulation groups.

To minimise confounding effects, the initiation of new treatments for NP and alterations to existing regimens will, in principle, be prohibited from the time of enrolment until week 8. Exceptions may be permitted when required to ensure participant safety.

### Outcomes

The primary outcome is the change in weekly average pain intensity scores recorded in a pain diary, using an 11-point NRS ranging from 0 (no pain) to 10 (worst imaginable pain). Participants will evaluate their average pain over the past 24 hours daily at home. Weekly averages will be calculated, and changes from the baseline week to each post-intervention week up to week 8 will be assessed. The primary endpoint is the change in weekly average pain scores from baseline to week 8.

Secondary outcomes comprise patient-reported and physiological measures, enabling a multidimensional evaluation of pain, functional impact, psychological status, neurophysiological response, and patient perception over time:

Pain NRS: Assessed separately from the diary, with participants reporting their current pain on the same 11-point scale.

SF-MPQ-2: Evaluates the qualitative aspects of pain, with participants rating 22 items based on their current pain experience [21].

BPI: Assesses pain presence, location, intensity, analgesic use, and interference with daily life across 16 self-report items [22].

EQ-5D-5L: Measures health-related quality of life across five domains [23].

HADS: Screens for anxiety and depression using 14 items [24].

PCS: Assesses maladaptive cognitive responses to pain [25, 26].

PSEQ: Measures confidence in maintaining function despite pain [27].

MMSE: Administered by assessors to screen for cognitive impairment.

QST: Conducted with thermal and vibration stimulators (TSA-II and VSA-3000, Medoc, Israel) based on published methods [32]. Warm, cold, heat pain, cold pain, and vibration detection thresholds will be assessed on the volar forearm or calf of the painful side. Each session includes two practice and four formal measurements. These assessments will be performed only when the stimulation target is the upper or lower limb.

Motor cortical excitability: Evaluated using single- and paired-pulse TMS with a figure-8 70-mm coil, two Magstim 200 stimulators, and a Bistim module (Magstim Company, Carmarthenshire, UK). MEPs will be recorded from the first dorsal interosseous muscle on the painful side (Brainsight MEP module, Rogue Research Inc.), elicited by contralateral M1 stimulation. Measures include resting motor threshold (RMT), short-interval intracortical inhibition, intracortical facilitation, and cortical silent period [33]. The motor hotspot is defined as the site eliciting the largest MEP, and RMT as the lowest stimulation intensity evoking MEPs ≥ 50 μV in at least five of 10 trials. Paired-pulse TMS will use a conditioning stimulus at 80% RMT and a test stimulus at 120% RMT, with interstimulus intervals of 2 ms (short-interval intracortical inhibition) and 15 ms (intracortical facilitation), plus control stimuli alone. Each stimulus will be delivered 10 times in a randomised order. The cortical silent period will be measured 10 times under 10–20% maximum voluntary contraction at 130% RMT, with an inter-trial interval of at least five seconds.

Paraesthesia (*shibire*) NRS: Participants will rate intensity on a 0–10 scale. “*Shibire*” is a common Japanese term referring to abnormal sensations such as paraesthesia, dysaesthesia, or numbness, and may also indicate motor difficulty, although it most often describes positive sensory symptoms such as paraesthesia [28].

*Shibire* Questionnaire [28]: Evaluates the presence, quality, location, duration, intensity, and daily life impact.

PGIC: Administered at week 8, with participants rating overall perception of change on a 7-point Likert scale (“very much improved” to “very much worse”).

Adverse events: Defined as any unfavourable or unintended sign, symptom, or illness occurring in a participant, regardless of causal relationship to the study. Worsening of pre-existing conditions will also be considered adverse events.

Blinding assessment: After the intervention, participants will be asked whether they believe they received real stimulation, sham stimulation, or are unsure.

### Statistical analysis

Efficacy analyses will be performed on the full analysis set, defined as all randomised participants who received at least one stimulation session and had at least one post-intervention outcome assessment (intention-to-treat analysis). A per-protocol set, excluding participants with major protocol deviations, will also be analysed for sensitivity. For participants with missing week 8 data, the last observation carried forward method will be applied to impute missing values for week 8 comparisons. Any modifications or additions to the analyses after study initiation will follow an evaluation of their validity and potential impact, with corresponding amendments to the statistical analysis plan. Handling of outliers will be pre-specified, and depending on the variable, either data transformation or statistical approaches robust to outliers will be applied.

Demographic characteristics and outcomes will be summarised descriptively, with longitudinal outcomes presented as line graphs over time. The primary analysis will use a linear mixed-effects model for repeated measures (MMRM) to evaluate changes in weekly average pain scores from the pain diary. The model will include change from baseline in weekly average pain scores as the response variable, with treatment group, time point, and their interaction as fixed effects, and individual participants as a random effect. Baseline pain scores will be included as covariates. Dunnett’s multiple comparison procedure will be used to assess statistically significant changes from baseline, and between-group differences will be explored at each time point. To assess clinically meaningful improvements, responder analyses will identify the proportion of participants achieving reductions of ≥2 points and ≥4 points [34] in weekly average pain scores at week 8 compared with baseline. Between-group comparisons will be performed using logistic regression models adjusted for baseline pain scores. Secondary outcomes measured repeatedly, such as pain NRS and SF-MPQ-2, will also be analysed using MMRM. Outcomes assessed only at baseline and week 8—including BPI, EQ-5D-5L, HADS, PCS, PSEQ, MMSE, QST, and MEP—will be evaluated using analysis of covariance, with change from baseline as the response variable, treatment group as a fixed effect, and baseline value as a covariate. Paraesthesia-related outcomes will be analysed using MMRM, analysis of covariance, or logistic regression, depending on data type and timing. PGIC responses at week 8 will be summarised by treatment group and compared using Fisher’s exact test. All analyses will be two-sided, with p-values < 0.05 considered statistically significant.

### Patient and public involvement

Patients and members of the public will not be involved in the design, recruitment, or conduct of this study.

## ETHICS AND DISSEMINATION

This study will be conducted in accordance with the ethical principles of the Declaration of Helsinki and the Clinical Trials Act of Japan. The study protocol was reviewed and approved by the University of Osaka Clinical Research Review Board (approval number S24007). All protocol amendments will be reviewed by the committee. Participants will be provided with detailed information about the study by the investigators, and written informed consent will be obtained before enrolment. Participation is voluntary, and consent may be withdrawn at any time. To protect confidentiality, each participant who provides informed consent will be assigned an identification code, ensuring that individuals cannot be directly identified. The correspondence table linking codes to personal information will be securely stored to prevent any disclosure of personal data outside the trial.

The clinical study report will be made publicly available via the jRCT website following review by the Clinical Research Review Board and the institutional administrator. Findings will also be disseminated through scientific presentations and peer-reviewed publications, with strict adherence to confidentiality. Data will be available from the corresponding author upon reasonable request.

## Supporting information

SPIRIT 2025 checklist

## Data Availability

Data will be available from the corresponding author upon reasonable request.

## ACKNOWLEDGEMENTS

English language editing was provided by Paperpal (www.paperpal.com) and Editage (www.editage.com).

## AUTHOR CONTRIBUTIONS

Hosomi K and Mori N contributed to the conceptualisation and methodology. Hoshikuma Y prepared the original draft. All authors contributed to review and editing. Kishima H supervised the study.

